# Conflicts of Interest in Cardiology Journals

**DOI:** 10.1101/2023.01.21.23284840

**Authors:** Karsha M Smith- Manga, Atiba B Manga, Brian J Piper

## Abstract

**Background:** Conflicts of interest are an ongoing concern in medical research. This takes place when the sponsor reports positive findings or promotes products over competitors because of their relationship and interaction with their industry sponsors. The *Physicians Payment Sunshine Act* mandates physicians who receive payments/compensation disclose their current and past relationships with different medical companies and the products they produce as an aim to manage conflicts of interest. In this quantitative bioethics study, we quantified financial conflicts of interest among cardiology journal authors and evaluated disparities in industry support among female and male physician-authors.

**Methods:** We reviewed 966 authors from 147 empirical research articles from two cardiology journals, *Circulation*, JCR 2021 Impact Factor (IF)= 29.7 and *Journal of the American College of Cardiology* (*JACC*), IF= 24.1, and one cardiovascular subspecialty journal, *Catheterization and Cardiovascular Interventions* (*CCI*), IF= 2.7. Articles published between January 1, 2020 and December 31, 2020 were reviewed. The database, OpenPayments.cms.gov (CMS-OP) was used to search author physician’s payments who reported receiving general payments, research payments, associated research funding, and ownership and investment interest between 2017 and 2019.

**Results:** A total of 19,529 payments totaled to 69,591,343.18 USD within the 36-month disclosure window. JACC accounted for 51.90%, Circulation 30.59% and CCI 17.51% of the total payment amounts. Male physician authors were more likely to receive industry sponsorship than their female counterparts (χ^2^ (1)= 23.30; *p*<0.00001). The 139 male physicians with CMS-OP accounts made up 90.23% of payments, while females accounted for 9.77%.

**Conclusion:** In conclusion, cardiology journal authors received appreciable renumeration form industry. Female cardiology authors had lower representation in authorship and honorarium compared to their male colleagues. There were also occasional author disclosure discrepancies, where some authors failed to report their relationship with financial institutions, despite each author receiving greater than one million dollars (USD) over a 36-month window. The evidence from this investigation supports that financial conflicts of interest is an ongoing issue in cardiology journals.

## Introduction

Conflicts of interest in cardiovascular medicine increase the risk of bias in healthcare judgment and medical research which is an ongoing concern in medical research.^1,2,3^ Financial conflicts of interest result when a person in a position of influence or power makes decisions based on compensation or other forms of personal gain.^4,5^ Authors who receive industry funding reported more positive research findings or subjects of their industry sponsors than those who reported receiving no financial sponsorship.^2, 6^ Concerns arise if research interests are influenced by secondary interests because of the clinical recommendations of certain medical interventions and procedures that are not as effective or safe as authors may lead them to be.^1, 2, 7^ This issue has been especially notable in cardiology journals as many research prospects rely on sponsorship to fund their investigations due to the increasing cost and complexity of research.^8^

The US *Physicians Payment Sunshine Act* mandates that payments for medical supplies, drug, biological, and medical device companies to disclose the relationships between physicians and their immediate families per Section 1128 G of the Social Security Act passed in 2010.^3^ Physicians who receive payments/compensation are requested by medical journals to disclose their current and past relationships with different medical companies and the products they produce. The Sunshine Medical Act may help manage conflicts of interest through disclosure.^3, 9^ and with public access to open payments on government websites like Centers for Medicaid and Medicare Services Open Payment program database (CMS-OP), one can make their own informed decisions on their overall health care.^10^ However, there are still limitations of payment disclosure. Underreporting of financial conflicts of interest in medicine is an ongoing concern.^11^

In this quantitative bioethics study, we investigated financial conflicts of interest in cardiology journals by looking at distinct payments that companies were paying to physicians to conduct research and promote their products over competitors. We also reviewed author disclosure statements to explore any discrepancies in physicians self-reporting. Finally, we examined disparities in industry earnings by gender. There are gender differences in salary among men and women in medicine.^12, 13^ and recent research suggests that industry payments are associated with the gap in revenue.^13, 14^

## Methods

### Procedures

For this research, 27 cardiology and cardiology subspecialty journals were initially screened based on their impact factor, whether they were published in the United States (US), and whether they were accessible for review. Two general cardiovascular journals were selected: *Circulation*, Journal Citation Report (JCR) 2021 Impact Factor (IF)= 29.69 and *Journal of the American College of Cardiology* (JACC), IF= 24.09; and one cardiovascular subspecialty journal: *Catheterization and Cardiovascular Interventions* (CCI), IF=2.69.

Empirical research articles from issues from December 31, 2020, proceeding in reverse chronological order to January 1, 2020, were collected, excluding systematic reviews or meta-analyses. Information extracted for authors of eligible articles included: author’s location, author’s first name, last name, middle initial, gender,^15^ educational degree, location, author position if they have an entry on CMS-OP, and the amount they received payments from 2017-2019. All author information, as previously described for the first ten eligible articles in *Circulation*, was collected in a pilot study to determine which author position was more likely to receive payments. Based on this information (Supplemental Figure 1), we selected the first three and last three authors for empirical research articles.

We collected payment information for the first 55 authors of each journal who received payments between 2017 and 2019 according to OpenPayments.cms.gov (CMS-OP) The payments include general payments, research payments, associated research funding, and ownership and investment interests. CMS-OP defines general payments as non-associated research payments, including compensation which may include transferred value for food and beverage, consulting fees, compensation for serving as a faculty or as a speaker for a non-accredited and non-certified continuing education program or medical education program, gifts, honoraria, travel and lodging, entertainment, charitable contributions, debt forgiveness, and current or prospective ownership or investment interest. CMS-OP defines associated research funding as funding for research projects or investigations in which the physician is considered a principal investigator. Procedures were approved by the IRB of Geisinger as exempt.

### Statistical Analysis

Each physician’s three-year total payment ($ USD) was collated based on the CMS-OP amounts between 2017 and 2019. We compiled the top ten highest-paid physicians of *Circulation, JACC*, and *CCI* for further investigation and statistical analysis. This data also included each physician’s gender, degree, location, and article topic. Those who had a CMS-OP account but received no payments (N= 25) between the 36-month disclosure window were considered outliers and not included in the analysis. We used GNU PSPP 1.4.1 to perform statistical analyses and GraphPad Prism 9 for figures. Chi-Square tests were used to compare categorical data. A p-value less than 0.05 was interpreted as statistically significant.

## Results

We examined a total of 966 authors (129 females) from 147 empirical research articles published in Circulation (n= 63), *JACC* (n= 44), and *CCI* (n= 40). All non-US authors and authors not holding an MD or DO degree were excluded (n = 642). Thus 324 US-based physicians remained. The first and last three physician-authors were included for a total of 165 who had qualifying payments of more than $10 (Figure 1). Fifteen physicians received payments and were authors of more than one article so the payments were only included once in the total amounts listed. Of the 324 US-based physicians, 79.63% had MD degrees only, 19.44% had an MD and additional graduate/professional degrees, and 0.93% were DOs.

**Figure 1:**
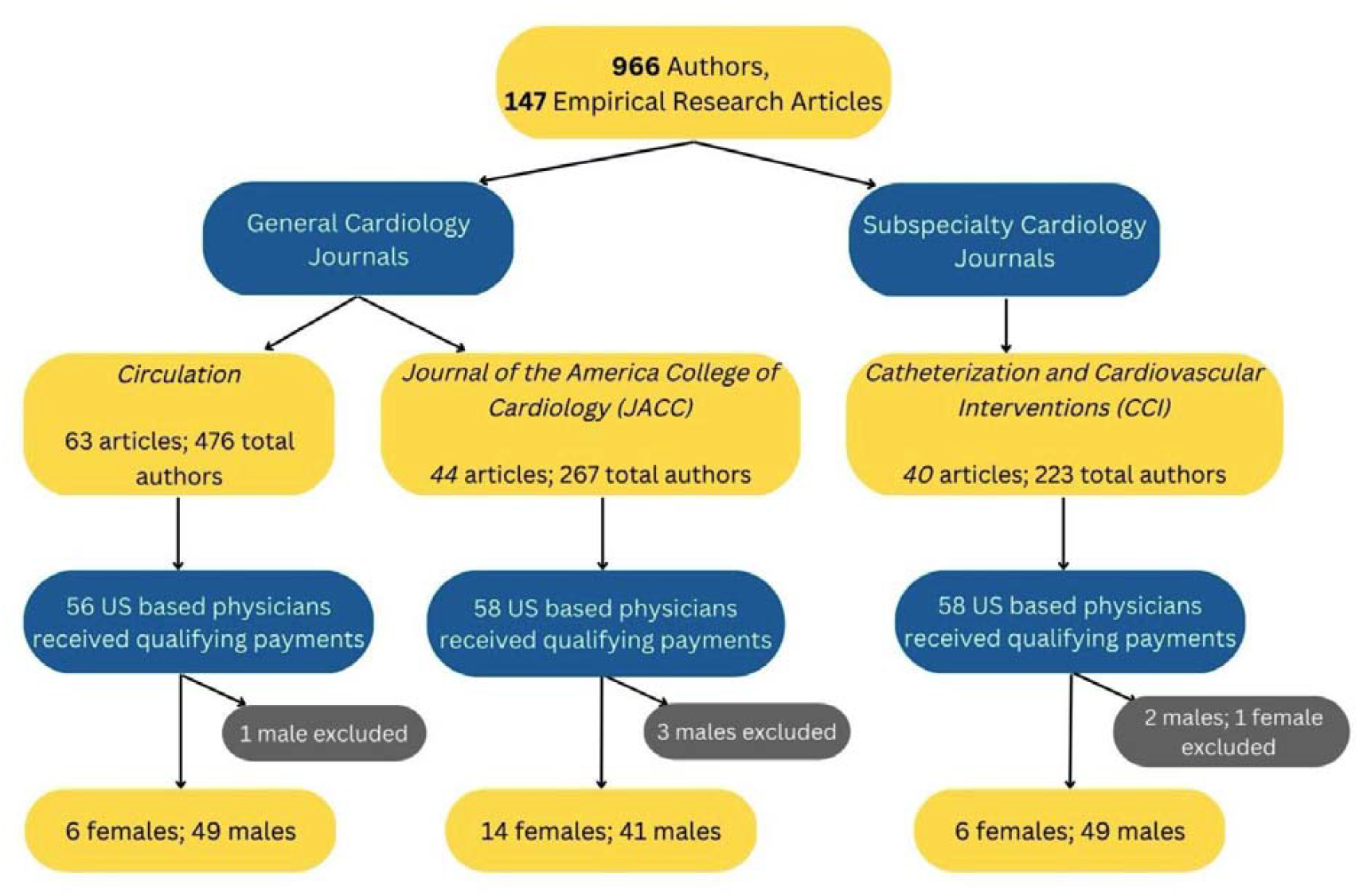
Flow diagram of screening cardiology journals for articles and authors based on author location and degree and their conflicts of interest as reported to the Centers for Medicare and Medicaid Services Open Payments. Second and additional entries of physicians were excluded if they authored more than one article and listed with the same author position.

Physician payments (19,529) totaled $69,591,343.18 within the 36-month disclosure window. *JACC* accounted for $36,118,859.34 (51.90%) from 7,676 payments across articles (63/147). *Circulation* articles (44/147) accounted for $21,289,241.05 USD (30.59%) over 6,661 payments, and *CCI* accounted for 17.51% of total payment amounts across articles (40/147). Of the total US authors, half (50.9%) had a qualifying CMS-OP entry. CCI had a similar percentage of authors with CMS-OP accounts (27.4%) compared to JACC (26.2%). *Circulation* constituted the lowest proportion of authors with CMS-OP (18.1%). The differences in CMS payments to physicians authored in *JACC* and Circulation were not statistically significant (*t*(108)= 1.3274; p=0.1872). However, CMS reimbursements to physicians authored in *JACC* and *CCI* was statistically significant (*t*(108)= 2.5924; p= 0.0109). Physician-authors were more likely to receive higher total payments if authored in *JACC* than in *CCI* (p= 0.03392, Figure 2B). Similarly, *Circulation* had higher payments than *CCI* (p= 0.00017, Figure 2B).

**Figure 2:**
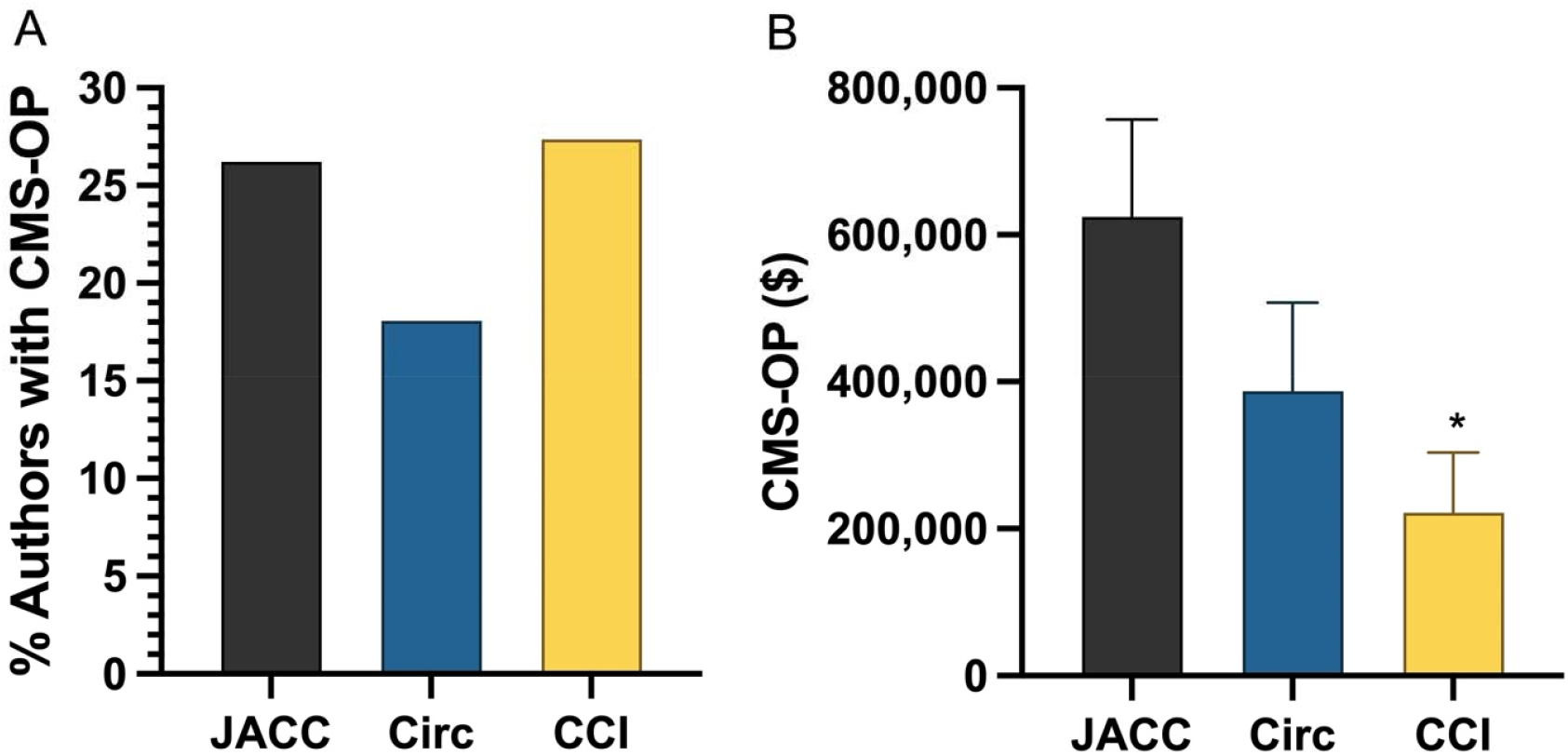
Percentage of authors with CMS-OP entries (A) and mean (+SEM) compensation (B) in *Circulation* (Circ), *Journal of the American College of Cardiology* (JACC), and *Catheterization and Cardiovascular Interventions* (CCI). *P< 0.05 vs JACC

**Figure 3:**
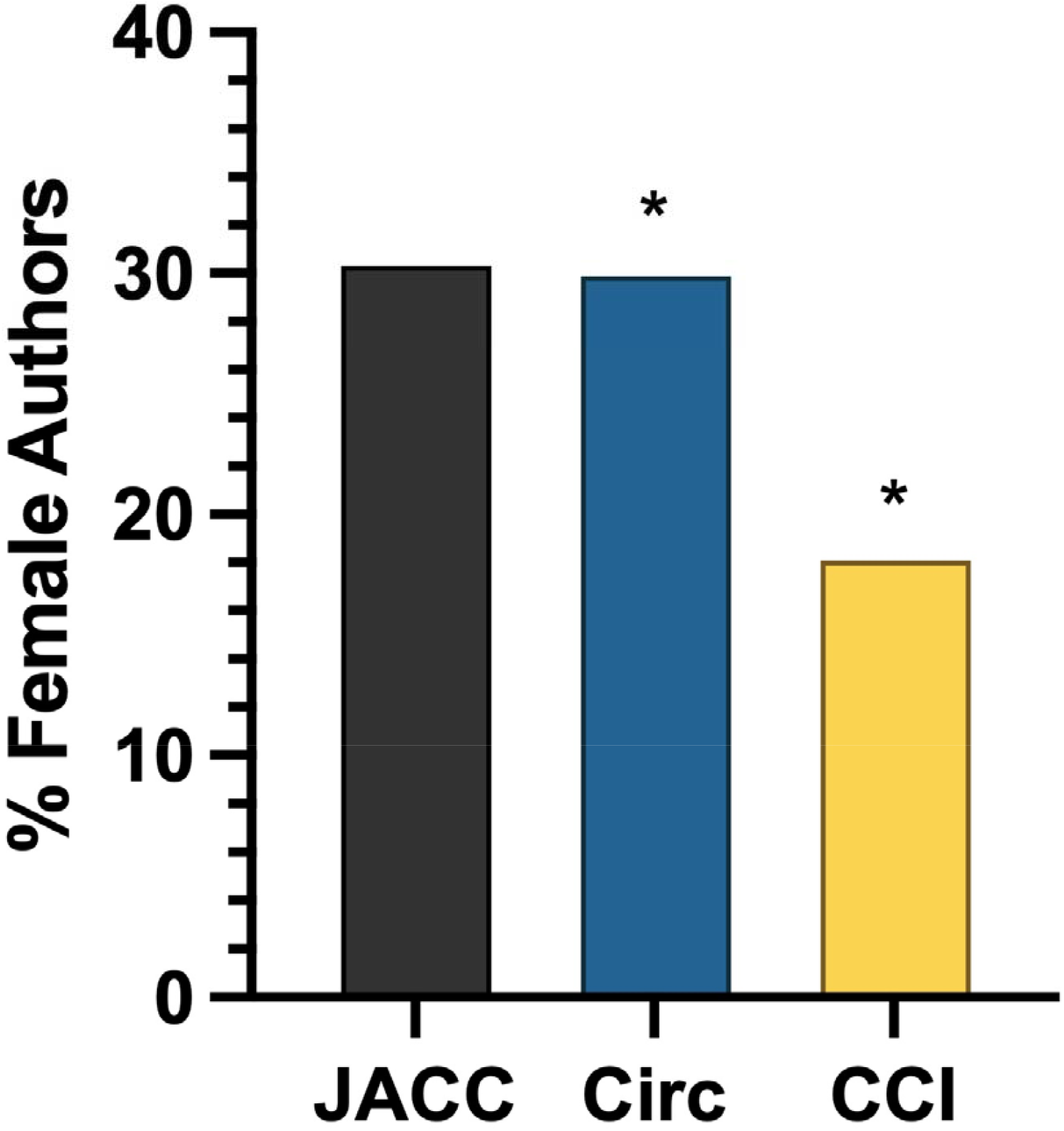
Percent of female authors in *Journal of the American College of Cardiology* (JACC), *Circulation* (Circ), and *Catheterization and Cardiovascular Interventions* (CCI). *JACC*= 30.30%, *Circ*= 29.88%, and *CCI*= 18.09%. *P< 0.05 versus JACC and Circulation

When comparing payments by gender, male physicians were significantly more likely to have a CMS-OP entry (16.7%) than their female colleagues (83.3.%, χ^2^(1) = 23.30; *p* < 0.00001). Overall, 139 male physician payments totaled to $62,791,804.44 whereas 26 female physician payments totaled to $6,799,538.74. *CCI* had the highest percentage of authors with CMS-OP accounts (27.35%) but had the fewest female authors with CMS-OP accounts (18.09%). *JACC* had the highest distribution of female authors with CMS-OP, 30.30%, and *Circulation* had 29.88% female authors with CMS-OP. One quarter (129/966 or 27.62%) of US-based authors were female. However, females only accounted for 9.77% of payments with a total of 26 female physicians with CMS-OP accounts. Conversely, the 139 male physicians with CMS-OP accounts made up for the preponderance (90.23%) of payments.

Physicians that reported having an MD degree and no additional degrees were significantly more likely to have a CMS-OP account than physicians who had MD and additional degrees (χ ^2^(1) = 4.28; p= 0.0385). Four-fifths (81.4%) of US-based physicians with CMS-OP accounts had MD degrees only which was more than four times the total of US-based physicians with MD degrees and additional degrees (PhD, MPH, MHS, MBBS, MBA, ScM, MHSc, MSME) at 17.21%. DO physicians accounted for 1.3% US-based physicians with an CMS-OP entry.

A chi-square analysis of CMS-OP entry to compare similarities among journals was completed. This analysis revealed χ ^2^= 31.8138; p <0.00001 for *CCI* vs *Circulation*. Similarly, *CCI* vs *JACC* was significant (χ ^2^= 4.4986; p= 0.033922) as was *JACC* vs *Circulation* (χ ^2^ (1)= 14.09 and p= 0.000174, Figure 2A).

When comparing payments for each journal, physicians receiving payments publishing in *JACC* had the highest mean value ($624,566.71, SEM: $132,155.45) with a median of $131,399.82. Physicians authored in *Circulation* had the second highest mean ($387,083.71, SEM: 120,600.43) with a median value of $66,077. *CCI*’s mean ($221,513.51, SEM: 81,902.14) and median ($19,154.83) were much lower. Three-fifths (59.4%) of physicians that received payments were the last three authors and two-fifths (40.6%) were the first three authors.

The top twenty highest compensated physician authors received 64.9% of the total remuneration. Among the twenty highest compensated (Table 1), 17 (85%) had MD degrees only, two had MD/PhD, and one MD/MPH. Two MD/PhD physician was among the top 20 highest paid authors with an aggregate of $3.954,051.50. There were two females with cumulative amount equaling $3,467,962.41 (7.7%). Two of the top twenty highest compensated reported no conflicts of interest in author disclosures. One of the highest paid authors from JACC reported being a non-paid member and reported in his disclosures receiving consultant fees only which accounts for 0.8% of their total remuneration over the 36-month disclosure window ($18,500).

**Table 1:** Top twenty highest conflicts of interest over 36-month disclosure window as reported to Centers for Medicare and Medicaid Services Open Payments among physicians-authors publishing in 2020 in three cardiology journals (JACC: *Journal of the American College of Cardiology; CCI: Catheterization and Cardiovascular Interventions)*. Author position = last (L), middle (M) or first (F).

| <u>Rank</u> | <u>Author Position</u> | <u>Degree(s)</u> | <u>Journal</u> | <u>Gender</u> | <u>Total 3Y Amount (USD)</u> | <u>Author Disclosure</u> |
| --- | --- | --- | --- | --- | --- | --- |
| 1 | L | MD | Circulation | M | 6,173,781.20 | Yes |
| 2 | M | MD | JACC | M | 4,435,834.08 | Yes |
| 3 | L | MD | CCI | M | 3,955,439.61 | Declared “none” |
| 4 | M | MD | JACC | M | 3,135,201.64 | Yes |
| 5 | M | MD/PhD | JACC | M | 2,896,093.50 | Yes |
| 6 | F | MD | JACC | M | 2,825,918.24 | Place of affiliation received grant, reports no personal financial relationship. |
| 7 | M | MD | JACC | M | 2,270,340.20 | Yes, consultant only |
| 8 | F | MD | JACC | M | 2,159,628.22 | Yes |
| 9 | M | MD | JACC | M | 1,887,655.89 | Yes |
| 10 | M | MD | CCI | F | 1,835,694.26 | Yes |
| 11 | M | MD | JACC | M | 1,768,635.37 | Yes |
| 12 | M | MD | Circulation | F | 1,632,268.15 | Yes |
| 13 | L | MD/MPH | Circulation | M | 1,600,223.82 | Yes |
| 14 | M | MD | JACC | M | 1,457,338.70 | Yes |
| 15 | F | MD | JACC | M | 1,343,296.86 | Yes |
| 16 | F | MD | JACC | M | 1,315,516.56 | Yes |
| 17 | M | MD | Circulation | M | 1,293,228.71 | Yes |
| 18 | M | MD | CCI | M | 1,137,993.92 | Yes |
| 19 | L | MD/PhD | JACC | M | 1,057,958.00 | Declared “none” |
| 20 | L | MD | Circulation | M | 962,446.26 | Yes |

## Discussion

There were three key findings in this investigation. This includes (1) discrepancies in the author’s disclosure, (1) the differences in total CMS-OP payments among *JACC, CCI*, and *Circulation*; and (3) the disparities in payments among genders. Industry remuneration to cardiology journal authors was appreciable ($69.6 million). When comparing all general and subspecialty cardiology journals, *JACC* and *Circulation* had the highest aggregate CMS-OP payments and average payments when compared to CCI. CCI was most likely to have authors that received payments, but on average had the least total payments compared to the non-subspecialty journals. Also noteworthy to mention, payments among the three highest-paid authors had higher amounts from medical device and supply manufacturers than pharmaceutical manufacturers. Furthermore, authors having MD degrees only were more likely to receive payments than their colleagues with MD and additional graduate degrees.

There were discrepancies in author disclosures, which is a frequent issue across many prior studies.^16, 17, 18, 19^ Four of the top 20 highest-paid authors who received sponsorship and honoraria from institutions, had discrepancies in their disclosures, with cumulative payment of $10,109,656. Author disclosure forms were created to show potential financial conflicts of interest; it also shows readers who are funding physician researchers.^19^ In this investigation, two of the twenty highest-paid authors reported no financial disclosures, one from JACC and CCI. The physician from CCI authored an article that investigated the efficacy of a transcatheter heart valve from a medical device manufacturer that issued payments to this author. The compensation from this specific company accounted for 10.2% of the author’s total amounts within the author’s 36-month disclosure window. The next physician from JACC who reported no disclosures authored an article studying the cardiovascular effects of patients post-surgical intervention. However, in his manuscript, there were no reviews on specific medical devices or drugs; lessening the concerns for bias. In addition, the sixth highest-paid author from JACC reported no personal financial affiliations despite CMS-OP listing payments to this physician. The physician was listed as the first author in an article investigating postoperative effects following mitral valve repairs with a device owned by a medical device manufacturer. The author reported positive outcomes for this device and the company accounted for 12% of remuneration over the 36-month disclosure window. Notably, the author did not receive payments in 2017 from this company which would then account for 95% over a 24-month disclosure. Lastly, the seventh highest-paid author reported only receiving compensation from consultations which only accounted for 0.8% of their remuneration over the three years. Author disclosures are especially vital in research to help notify readers of potential bias over studies. In addition, author disclosures serve to decrease the risks of biased research findings when sponsorship and funding are revealed.^20^ Prior studies show physician researchers are more likely to publish results that are advantageous to their sponsors or industries from which they receive compensation.^18^ These findings further support the evidence of conflicts of interest and underreporting in author disclosures.

Another finding concerning for conflicts of interest is the payments among female and male physicians. Female authorship in all journals were roughly similar, ranging from 26% to 30%. Women account for the majority of medical school students in DO and MD granting institutions, but an existential gap remains when it comes to gender equity among physicians where women only account for 13% of US practicing cardiologists.^21^ There were significant differences in conflicts of interest among male and female physician-authors. Female authors made up more than one-fourth of the total authors, but only accounted for one-tenth of the total CMS-OP payments. Out of the top 10 highest-paid physicians, one was a female and she had the lowest total amount of CMS-OP remuneration. The data and gaps between female and male physician authors seen in this investigation support the evidence of underrepresentation of female authorship in cardiology scientific articles.^22, 23^

Historically, the distribution of women is also lagging in academic medicine, which is an avenue for career advancement.^24^ Because of this, mentorship and sponsorship were a potential solution for leadership diversity, but women also lack sponsorships in academic medicine compared to their male colleagues.^25^ Some research states this is due to the characteristics of mentors or the number of sponsors available to women.^22, 25, 26^ Sponsorship has been shown to have a positive impact on the advancement of one’s career ^25,27^ and the increased credibility for women rely on additional support from sponsors.^24^ While there has been a modest growth of female physicians in medicine; workplace gender disparities remain and affect women’s growth in their careers. This happens because of the limitations on sponsorship and placement in high-level executive positions in academic medicine.

Some limitations of this study include the limited sample pool. Assessing only the first three and three senior authors of each article based on seniority may not collectively represent all authors. However, we completed a pilot study on the first 20 articles in *Circulation*. The data showed that out of 41 authors who received CMS-OP disbursements greater than 10 USD, 20 were among the first or last three in authorship. The remaining 21 authors were skewed in the author position and only 2 were median authors. Second, there was relatively infrequent data on the female authors in this study. Although this reflects the current status of the field, interpretation of gender differences should be made with caution. Finally, other potential conflicts of interest are not quantifiable. The Physician Payment Sunshine Act only mandates payments from manufacturers that were reimbursed by Medicare, Children’s Health Insurance Program, and Medicaid to be reported, whereas manufacturers who do not receive these reimbursements are not required to report payments distributed to physicians.^18^

In summary, over half (50.9%) of US-based physician-authors publishing in three cardiology journals received CMS-OP reported payments of $69.6 million. Male physicians were more likely to be featured as first or senior authors and receive higher total payments than their female colleagues, which brings about concerns for conflicts of interest. It is important to acknowledge some research issues are more likely to encourage funding and sponsorship.^5^ Preferences over certain research topics also pose a risk for conflicts of interest and therefore a lack of funding on topics with less focus. Quantitative research of conflicts of interest in cardiology will continue to be an existential concern in scientific research and future studies will be important in informing solutions to maintaining the highest standards for trustworthy evidence-based medicine. Future investigations should also focus on what factors contribute to disparities in industry payments across genders and if physician authorship in certain journals are more likely to bring about higher industry remuneration.

## Data Availability

All data produced in the present study are available upon reasonable request to the authors.

## Acknowledgements

Stephen Voyce and Gabi Waite provided valuable information and insight. Iris Johnston provided valuable support.

