## Supplemental Figures for "Conflicts of Interest in Cardiology Journals"

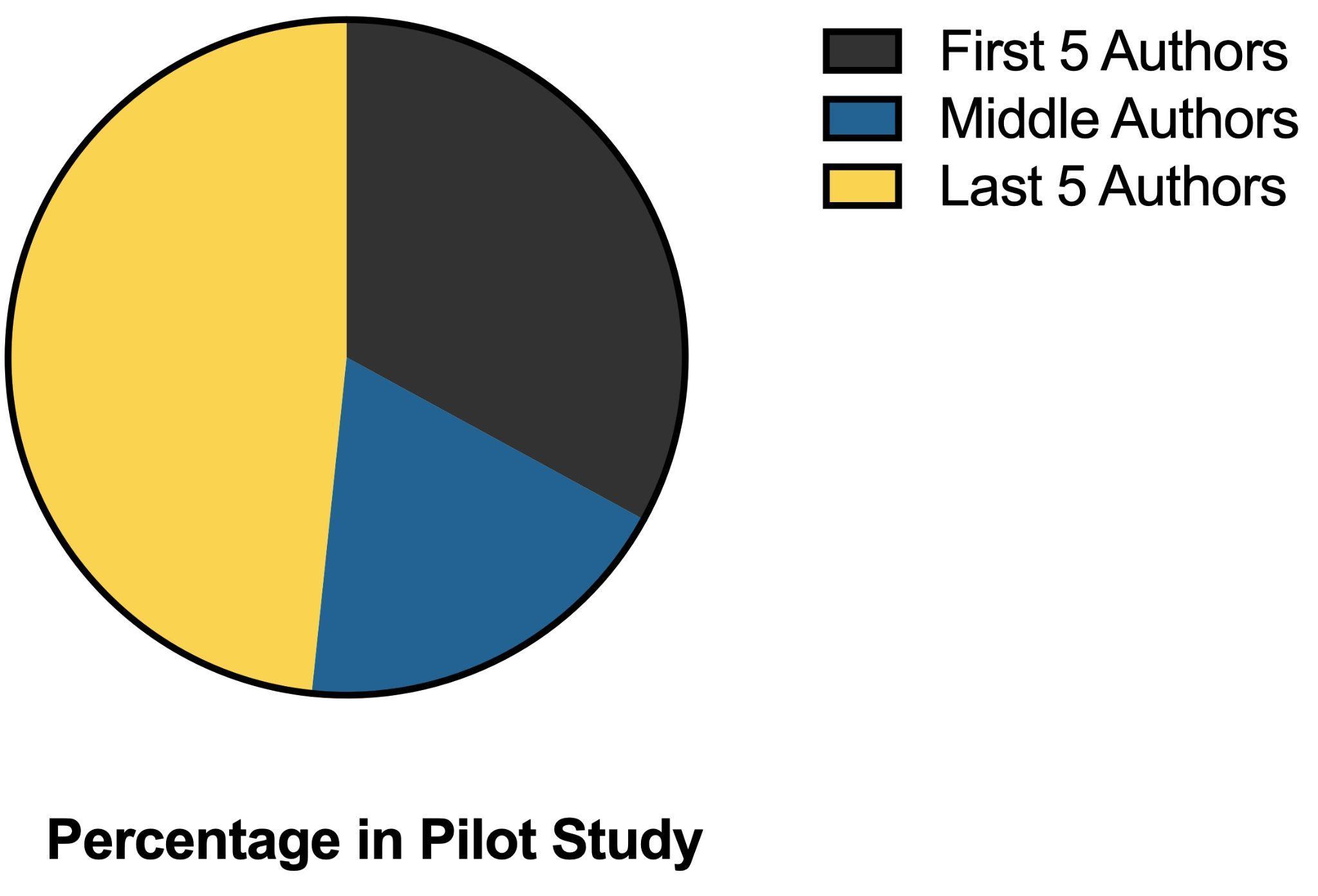

**Supplemental Figure 1:** Percent of total compensation ($9,869,096.89) by author position in first 20 articles of *Circulation*. First five authors= 33.3%, Non-first/last authors= 17.1%, Last five authors= 44.4%.

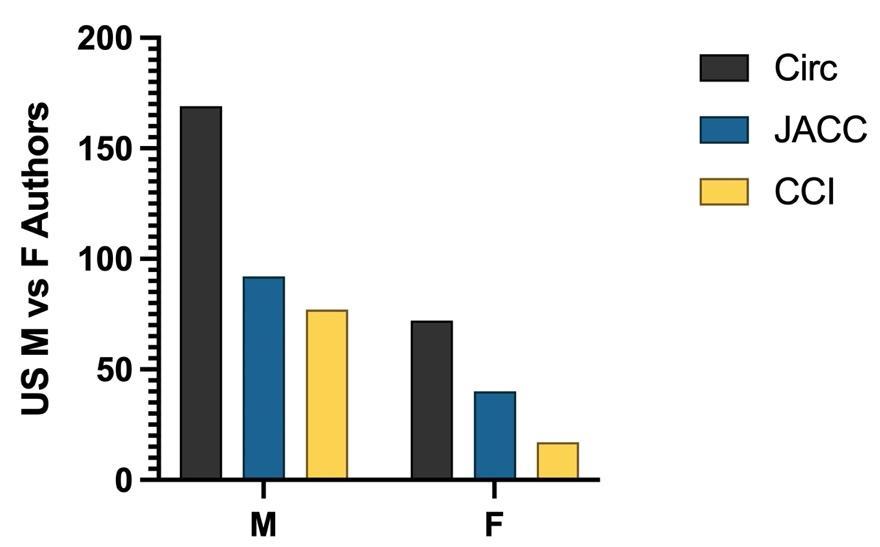

**Supplemental Figure 2:** Total number of male (M) and female (F) authors in *Circulation* (Circ), *Journal of the American College of Cardiology* (JACC), and *Catheterization and Cardiovascular Interventions* (CCI). *Circ* M=169 and F= 72. *JACC* M=92 and F= 40. *CCI* M=77 and F=17

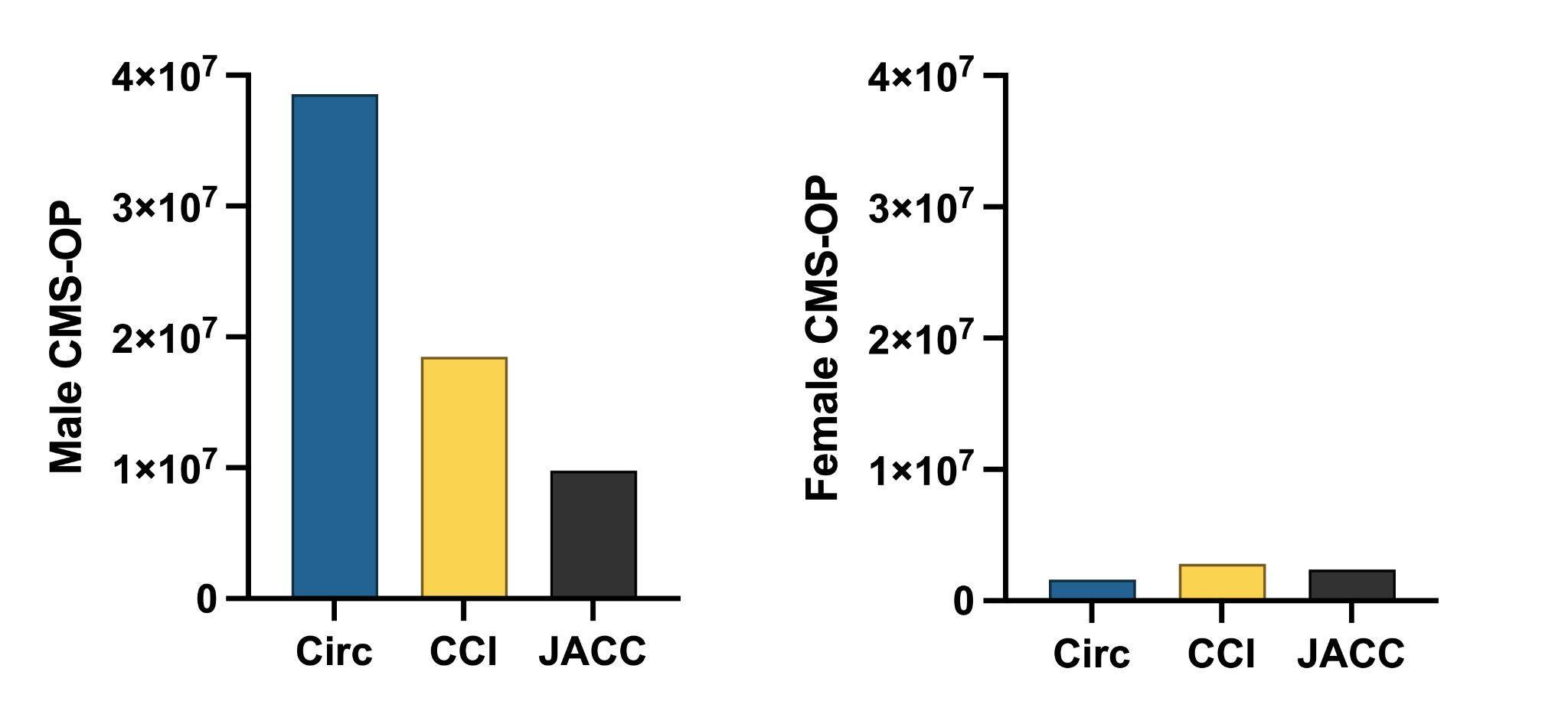

**Supplemental Figure 3:** Total CMS-OP compensation for male (left) and female (right) physician authors in *Circulation* (Circ), *Journal of the American College of Cardiology* (JACC), and *Catheterization and Cardiovascular Interventions* (CCI).

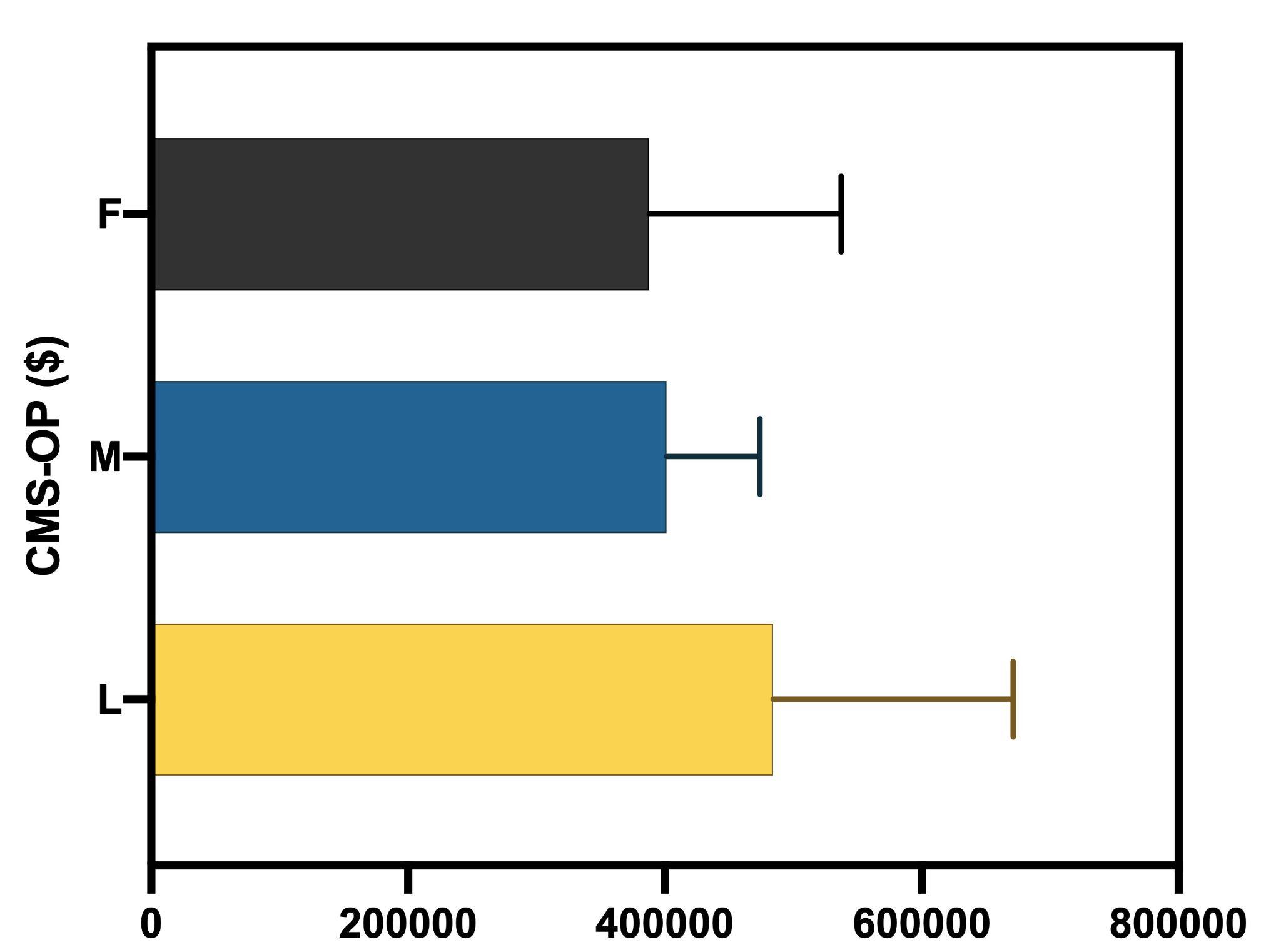

**Supplemental Figure 4:** Distribution of total CMS-OP payments for first and last three authors in *Circulation* (Circ), *Journal of the American College of Cardiology* (JACC), and *Catheterization and Cardiovascular Interventions* (CCI). (F=26, M=100, L=39)
